# Group A streptococcal cases and treatments during the COVID-19 pandemic and 2022 outbreak: a retrospective cohort study in England using OpenSAFELY-TPP

**DOI:** 10.1101/2023.09.22.23295850

**Authors:** Christine Cunningham, Louis Fisher, Christopher Wood, The OpenSAFELY Collaborative, Victoria Speed, Andrew D Brown, Helen J Curtis, Rose Higgins, Richard Croker, Ben FC Butler-Cole, David Evans, Peter Inglesby, Iain Dillingham, Sebastian CJ Bacon, Elizabeth Beech, Kieran Hand, Simon Davy, Tom Ward, George Hickman, Lucy Bridges, Thomas O’Dwyer, Steven Maude, Rebecca M Smith, Amir Mehrkar, Liam C Hart, Chris Bates, Jonathan Cockburn, John Parry, Frank Hester, Sam Harper, Ben Goldacre, Brian MacKenna

**Author notes:** **Abbreviations** GAS, iGAS, SSP, RR, UKHSA, EHR, IMD.

## Abstract

**Objective:** To investigate the impact of the COVID-19 pandemic on Group A streptococcal (GAS) cases and related antibiotic prescriptions.

**Design:** A retrospective cohort study with supporting dashboards with the approval of NHS England.

**Setting:** Primary care practices in England using TPP SystmOne software from January 2018 through March 2023.

**Participants:** Patients included were those registered at a TPP practice for each month of the study period. Patients with missing sex or age were excluded, resulting in a population of 23,816,470 in January 2018, increasing to 25,541,940 by March 2023.

**Main outcome measures:** We calculated monthly counts and crude rates of GAS cases (sore throat/tonsillitis, scarlet fever, invasive group A strep) and prescriptions linked with a GAS case, before (pre-April 2020), during and after (post-April 2021) COVID-19 restrictions. We calculated the maximum and minimum count and rate for each season (years running September-August), and the rate ratio (RR) of the 2022/23 season to the last comparably high season (2017/18).

**Results:** Recording of GAS cases and antibiotic prescription linked with a GAS case peaked in December 2022, higher than the 2017/2018 peak. The peak rate of monthly sore throat/tonsillitis (possible group A strep throat) recording was 5.33 per 1,000 (RR 2022/23 versus 2017/18 1.39 (CI: 1.38 to 1.40)). Scarlet fever recording peaked at 0.51 per 1,000 (RR 2.68 (CI: 2.59 to 2.77)), and invasive group A streptococcal infection (iGAS) at 0.01 per 1,000 (RR 4.37 (CI: 2.94 to 6.48)). First line antibiotics with a record of a GAS infection peaked at 2.80 per 1,000 (RR 1.37 (CI:1.35 to 1.38)), alternative antibiotics at 2.03 per 1,000 (RR 2.30 (CI:2.26 to 2.34)), and reserved antibiotics at 0.09 per 1,000 (RR 2.42 (CI:2.24 to 2.61)). For individual antibiotics, azithromycin with GAS indication showed the greatest relative increase (RR 7.37 (CI:6.22 to 8.74)).This followed a sharp drop in recording of cases and associated prescriptions during the period of COVID-19 restrictions where the maximum count and rates were lower than any pre COVID-19 minimum.

More detailed demographic breakdowns can be found in our regularly updated dashboard report.

**Conclusions:** Rates of scarlet fever, sore throat/tonsillitis and iGAS recording and associated antibiotic prescribing peaked in December 2022. Primary care data can supplement existing infectious disease surveillance through linkages with relevant prescribing data and detailed clinical and demographic subgroups.

**What is known:** During the COVID-19 pandemic there has been a substantial change to the pattern of circulating viruses and bacteria that cause illnesses. A spike in group A streptococcal infections in England starting December 2022 was associated with 426 deaths, including 48 children as of 7th May 2023. Increased demand for antibiotics in this period led to medicines shortages and the introduction of Serious Shortage Protocols (SSPs). Existing surveillance systems such as notifiable disease reports and GP in-hours surveillance bulletins describe clinical events, but they do not link to relevant prescribing data.

**What this study adds:**

- This study supports the findings of routine surveillance reports which indicated a drop in GAS infections during the COVID-19 restrictions, followed by a spike in December 2022, demonstrating that the OpenSAFELY platform and primary care data can be used to rapidly describe not only clinical events but also relevant prescribing in the case of future outbreaks.
- Antibiotic prescribing with a GAS indication, particularly for phenoxymethylpenicillin alternatives and reserved antibiotics, was higher in the December 2022 peak than in the 2017/2018 peak.

## Background

During the COVID-19 pandemic there has been a substantial change to the pattern of circulating viruses and bacteria that cause illnesses [1]. On December 2nd 2022, the UK Health Security Agency (UKHSA) issued an alert of higher than normal events of scarlet fever and other group A streptococcal (GAS) infections. Then on December 5th 2022 the UKHSA issued an urgent public health message to healthcare professionals advising a lower threshold for prescribing antibiotics to children presenting with features of GAS [2]. GAS infections are normally mild, often causing non-specific symptoms such as sore throat [3], or, with scarlet fever, more specific symptoms including “strawberry tongue” and a distinctive rash [4]. On rare occasions these bacteria can enter the bloodstream or deep tissues causing invasive group A streptococcal infections (iGAS), a serious infection which can lead to death. Between 19th September 2022 and 7th May 2023, there were 426 deaths associated with iGAS in England, including 48 children under 18 [5]. UKHSA indicates that the increase is likely to reflect increased susceptibility to these infections in children due to low numbers of cases during the COVID-19 pandemic, along with increased circulation of respiratory viruses [1]. Prior to this, 2017/18 was the last time significantly high case numbers were reported [5].

Phenoxymethylpenicillin is recommended as the first line treatment for scarlet fever, with clarithromycin as an alternative in penicillin allergic patients [6]. Following the UKHSA messaging in December 2022, it was anticipated that increased demand may lead to shortages of these drugs. Interim treatment guidelines were produced by the NHS England GAS Clinical Reference Group and the UKHSA Incident Management Team [7,8], and a series of Serious Shortage Protocols (SSPs) [9] were introduced, both recommending a range of alternative antibiotics, and the SSPs allowing community pharmacists to substitute an alternative antibiotic where the prescribed antibiotic was unavailable.

OpenSAFELY is a secure health analytics platform that allows near real-time analysis of pseudonymised primary care patient records in England. We have previously shown that the OpenSAFELY platform can be used for rapid audit and feedback for national health incidents [10]. In order to support the response we therefore set out to 1) conduct a rapid analysis to describe the recording of diagnoses and symptoms related to GAS infection, 2) assess the prescribing of a wide range of antibiotics potentially used to treat GAS infection, 3) give detailed contextual information for key patient groups, and 4) develop a dashboard for use by NHS and UKHSA staff. This is a framework that can be reused or repurposed for future outbreaks.

## Methods

### Study Design

With the approval of NHS England, we performed a retrospective cohort study of GAS and prescribed antibiotics on a monthly basis from January 2018 through March 2023. From September 2022 through March 2023 data was also extracted on a weekly basis to provide real-time weekly results as part of the rapid surveillance report. Patients were included in a monthly or weekly data extract if they were alive and registered at a TPP practice in that time period. Patients with a missing sex or age (those with age below 0 or above 120 years) were excluded.

### Data Sources and processing

All data were linked, stored and analysed securely within the OpenSAFELY platform: https://opensafely.org/. Data include pseudonymised data such as coded diagnoses, medications and physiological parameters. No free text data are included. All code is shared openly for review and re-use under MIT open licence here https://github.com/opensafely/strepa_scarlet/. Detailed pseudonymised patient data is potentially re-identifiable and therefore not shared. Aggregated data used in the analyses are available upon reasonable request to the authors. Data management and analysis was performed using Python 3.

### Identification of clinical events

Based on the recommendation given by NHS England, UKHSA interim clinical guidance [7,8] and the phenoxymethylpenicillin SSPs [9], we generated dm+d (dictionary of medicines and devices) codelists for the oral formulations of the following antibiotics: phenoxymethylpenicillin, amoxicillin, clarithromycin, erythromycin, azithromycin, flucloxacillin, cefalexin and co-amoxiclav. Antibiotics were additionally categorised into three groups: phenoxymethylpenicillin (Group 1) as the first line antibiotic for GAS infection; macrolides, amoxicillin and flucloxacillin (Group 2) as the antibiotics recommended in case of non-availability or penicillin allergy; and cefalexin and co-amoxiclav (Group 3) as the least preferred broad-spectrum antibiotics due to their greater risk of promoting antimicrobial resistance.

We also developed three SNOMED-CT codelists to identify the different types of GAS or possible GAS infection: 1) sore throat/tonsillitis (possible group A strep throat), 2) scarlet fever, and 3) iGAS. In the absence of a clear indicator of group A strep throat, we took a pragmatic approach whereby codes for sore throat/pharyngitis/tonsillitis have been included where an alternative cause for the sore throat is not stated (e.g.Staphylococcal tonsillitis), but non GAS cases may still be included. Scarlet fever and iGAS have clearer diagnosis codes, but as iGAS cases are more likely to occur in hospital than primary care, some cases may be missing from this analysis. All three codelists were included to provide a sensitive measure more likely to provide an early signal of changes in GAS case counts.

The medication and clinical codelists used in this analysis are shown in Supplemental Table S1 and are openly available for inspection and re-use on https://www.opencodelists.org/. Prescribing data is based on prescriptions issued within the Electronic Health Record (EHR), and does not necessarily reflect whether the prescription was dispensed. Clinical events data is based on a clinical code being added to a patient’s record. This is often added by a clinician during a consultation to indicate the presence of a sign/symptom (e.g. sore throat) or that a clinical diagnosis has been made (e.g. scarlet fever). These codes do not necessarily indicate positive test results.

### Outcomes

#### GAS Diagnoses and Symptoms

For each time period, we identified patients with clinical codes for each of sore throat/tonsillitis, scarlet fever, or iGAS. A separate measure was computed of patients with any of the above codes. In the supporting dashboards, we counted the number of patients with each clinical event (scarlet fever, sore throat/tonsillitis, or iGAS) who also had an antibiotic prescription up to 7 days prior or 14 days after the clinical event.

#### Antibiotics used to treat GAS infection

For each time period we also identified patients who had a record of each individual antibiotic and additionally counted patients with any antibiotic. We then grouped the antibiotics into Group 1 (first line), Group 2 (alternative), and Group 3 (reserved broad spectrum). Although all the antibiotics in this study can be used for GAS infection, they are not exclusively used for GAS infection. Therefore, the results focus on antibiotic prescribing in combination with any of the potential GAS clinical events of interest up to 14 days prior or 7 days after the prescribing event. Details on antibiotics with or without GAS indication can be found in the supplemental and dashboards.

#### Breakdowns by Demographic Subgroups

We extracted the following patient demographic information: age band in years (0-4, 5-9, 10-14, 15-44, 45-64, 65-74, and 75+), sex, 2019 Index of Multiple Deprivation (IMD) based on patient address as quintiles, ethnicity according to the 2001 census (White, Mixed, Asian or Asian British, Black or Black British, Chinese or Other, or Unknown), and the region of the patient’s practice based on the 9 Nomenclature of Territorial Units for Statistics (NUTS) regions for England (North East, North West, Yorkshire and The Humber, East Midlands, West Midlands, East, London, South East, and South West).

Demographic breakdowns for all outcomes are available in the supporting dashboards, but phenoxymethylpenicillin with GAS indication broken down by demographic group is presented in this paper.

Those with “Missing” values in each demographic group were excluded from the plots, but described in Table 1. The OpenSAFELY-TPP population has been shown to be broadly representative of the English population, but there are some regional coverage differences due to EHR system use, with the highest coverage in the East of England (91%) and lower coverage in London (19%) [11]. Care should be taken when interpreting regional rates, as they could reflect other demographic differences.

**Table 1:** Characteristics of registered patients and rates of sore throat/tonsillitis, scarlet fever, or invasive group A strep at the end of the study period (March 2023)

|  |  | Overall Population<br>No. registered patients (%) | Any GAS Clinical Event<br>Rate per 1,000 (95% CI) |
| --- | --- | --- | --- |
| Total |  | 25,541,940 (100.0) | 3.36 (3.34 to 3.38) |
| Sex | F | 12,748,390 (49.9) | 4.09 (4.06 to 4.13) |
|  | M | 12,793,540 (50.1) | 2.63 (2.60 to 2.65) |
| Region | East | 5,857,920 (22.9) | 3.73 (3.68 to 3.78) |
|  | East Midlands | 4,447,350 (17.4) | 3.36 (3.31 to 3.42) |
|  | London | 1,819,560 (7.1) | 2.60 (2.53 to 2.67) |
|  | North East | 1,189,140 (4.7) | 3.43 (3.33 to 3.54) |
|  | North West | 2,209,000 (8.6) | 3.92 (3.84 to 4.01) |
|  | South East | 1,656,800 (6.5) | 2.88 (2.80 to 2.96) |
|  | South West | 3,532,570 (13.8) | 2.59 (2.54 to 2.65) |
|  | West Midlands | 1,035,480 (4.1) | 3.85 (3.73 to 3.97) |
|  | Yorkshire And The Humber | 3,705,650 (14.5) | 3.58 (3.52 to 3.64) |
|  | Missing | 88,460 (0.3) | 3.73 (3.33 to 4.13) |
| Imd | 1 - Most Deprived | 5,044,510 (19.7) | 4.05 (4.00 to 4.11) |
|  | 2 | 4,928,760 (19.3) | 3.45 (3.40 to 3.50) |
|  | 3 | 5,231,200 (20.5) | 3.12 (3.07 to 3.17) |
|  | 4 | 4,909,180 (19.2) | 2.97 (2.92 to 3.02) |
|  | 5 - Least Deprived | 4,516,700 (17.7) | 2.81 (2.76 to 2.86) |
|  | Missing | 911,590 (3.6) | 5.21 (5.06 to 5.36) |
| Ethnicity | Black | 708,560 (2.8) | 3.02 (2.89 to 3.15) |
|  | Mixed | 492,010 (1.9) | 5.00 (4.80 to 5.20) |
|  | Other | 621,040 (2.4) | 3.01 (2.87 to 3.15) |
|  | South Asian | 2,064,330 (8.1) | 4.31 (4.22 to 4.40) |
|  | White | 19,669,920 (77.0) | 3.38 (3.35 to 3.40) |
|  | Missing | 1,986,080 (7.8) | 2.03 (1.97 to 2.10) |
| Age Band | 0-4 | 1,199,240 (4.7) | 11.85 (11.66 to 12.04) |
|  | 5-9 | 1,433,440 (5.6) | 11.07 (10.90 to 11.24) |
|  | 10-14 | 1,537,990 (6.0) | 5.68 (5.56 to 5.80) |
|  | 15-44 | 9,996,000 (39.1) | 3.64 (3.60 to 3.67) |
|  | 45-64 | 6,549,540 (25.6) | 1.18 (1.15 to 1.21) |
|  | 65-74 | 2,465,340 (9.7) | 0.73 (0.69 to 0.76) |
|  | 75+ | 2,360,370 (9.2) | 0.47 (0.44 to 0.49) |

**IMD = index of multiple deprivation**
**Patients with more than one of the same clinical event in a time period are only counted once** **Counts have been rounded to the nearest 10 and the rate computed with the rounded numbers** **Sums of category counts may not exactly match the "Total" due to rounding**

#### Statistical Methods

For each time period (month or week), cases and prescriptions were described with simple descriptive statistics of counts and crude rates. Counts represent patients with at least one prescription or clinical event in that time period. Patients with more than one of the same prescription or clinical event in a time period were only counted once. Counts <=5 have been redacted and all numbers rounded to the nearest 10 to avoid potential re-identification of patients. To compute the crude rate, the rounded count was divided by the included study population and multiplied by 1,000 to achieve a rate per 1,000 registered patients. All rates were calculated with 95% confidence intervals. Rates are used to account for the registered OpenSAFELY-TPP population increasing over time (Figure S1).

We compute the maximum and minimum count and rate of each GAS season (September through August) [5], and compare the pre-COVID-19 restriction years to the years with COVID-19 restrictions and the post COVID-19 restriction years. The COVID-19 restriction is defined as 1st April 2020 to 1st April 2021, when non-essential services began re-opening [12]. The rate ratio (RR) of the 2022/23 season maximum to the maximum of the last comparably high season (2017/18) was also calculated.

#### Dashboard

Regular dashboards have been produced and were routinely updated at https://reports.opensafely.org/ with near real-time data from January to April 2023. These dashboards contain separate sections for each type of GAS infection and antibiotic, as well as additional graphs. An example of an additional plot is top codes over time. Each codelist is composed of a number of codes. Scarlet fever, for example, can be coded in the patient’s medical record with the SNOMED-CT code 1087781000000109 “Suspected Scarlet Fever”, 170523002 “Notification of Scarlet Fever” and so on. The number of times each code was used was summed across the entire study period, and the count and proportion of the top 5 most commonly used codes was reported. The top 5 codes in the first month and the top 5 codes in the last month were then plotted over the entire study period.

#### Patient and public involvement

Patients were not formally involved in developing this study design. We have developed a publicly available website https://opensafely.org/ through which we invite any patient or member of the public to contact us regarding this study or the broader OpenSAFELY project.

## Results

At the start of the study period there were 23,816,470 alive and registered patients in OpenSAFELY-TPP with non-missing age and sex, which increased to 25,541,940 at the end of the study period in March 2023 (Table 1). This represents about 40% of the total patients registered at a practice in England [13,14]. In March 2023, 3,650 patients had a record of scarlet fever, 83,080 sore throat/tonsillitis, and 70 iGAS, affecting 85,800 (3.4 per 1,000) patients in total. Clinical codes for these infections were most common among ages 0-4 (11.8 per 1,000) and 5-9 (11.1 per 1,000). Females had a higher rate than males (4.1 per 1,000 vs 2.6 per 1,000), and clinical events increased with deprivation, ranging from 2.8 per 1,000 in the least deprived quintile to 4.1 per 1,000 in the most deprived quintile. The South West, London, and the South East had the lowest crude rates of clinical events (2.6, 2.6, and 2.9 per 1,000). The highest rate of clinical events across ethnic groups was in those with Mixed ethnicity (5.0 per 1,000), followed by South Asian (4.3 per 1,000), White (3.4 per 1,000), Black (3.0 per 1,000), and Other (3.0 per 1,000).

### Trends in GAS Diagnoses and Symptoms

In the three GAS seasons (2017/18, 2018/19, and 2019/20) prior to the COVID-19 restrictions, the number of sore throat/tonsillitis cases per month peaked each winter, at 91,510 (January 2018), 75,070 (January 2019) and 77,470 (December 2019) (Figure 1). During the 2020/21 year, which included the COVID-19 restrictions (April 2020 through March 2021), the typical winter peak was not observed, with a maximum count of only 30,860 in September 2020, lower than the minimum in the previous seasons (e.g. August 2018 (40,130) and August 2019 (41,000)). The count increased over 2021/22 but still without a distinct seasonal pattern. Starting in September 2022, there was a steep increase in events, peaking in December 2022 at 135,860, 44,350 higher than the previous highest count (January 2018), a rate ratio of 1.39 (CI:1.38 to 1.40) (Table S2). 75% of these sore throat/tonsillitis cases also had an antibiotic prescription up to 7 days prior or 14 days after the clinical event (Table S7), but they still may still include non-GAS cases.

**Figure 1:**
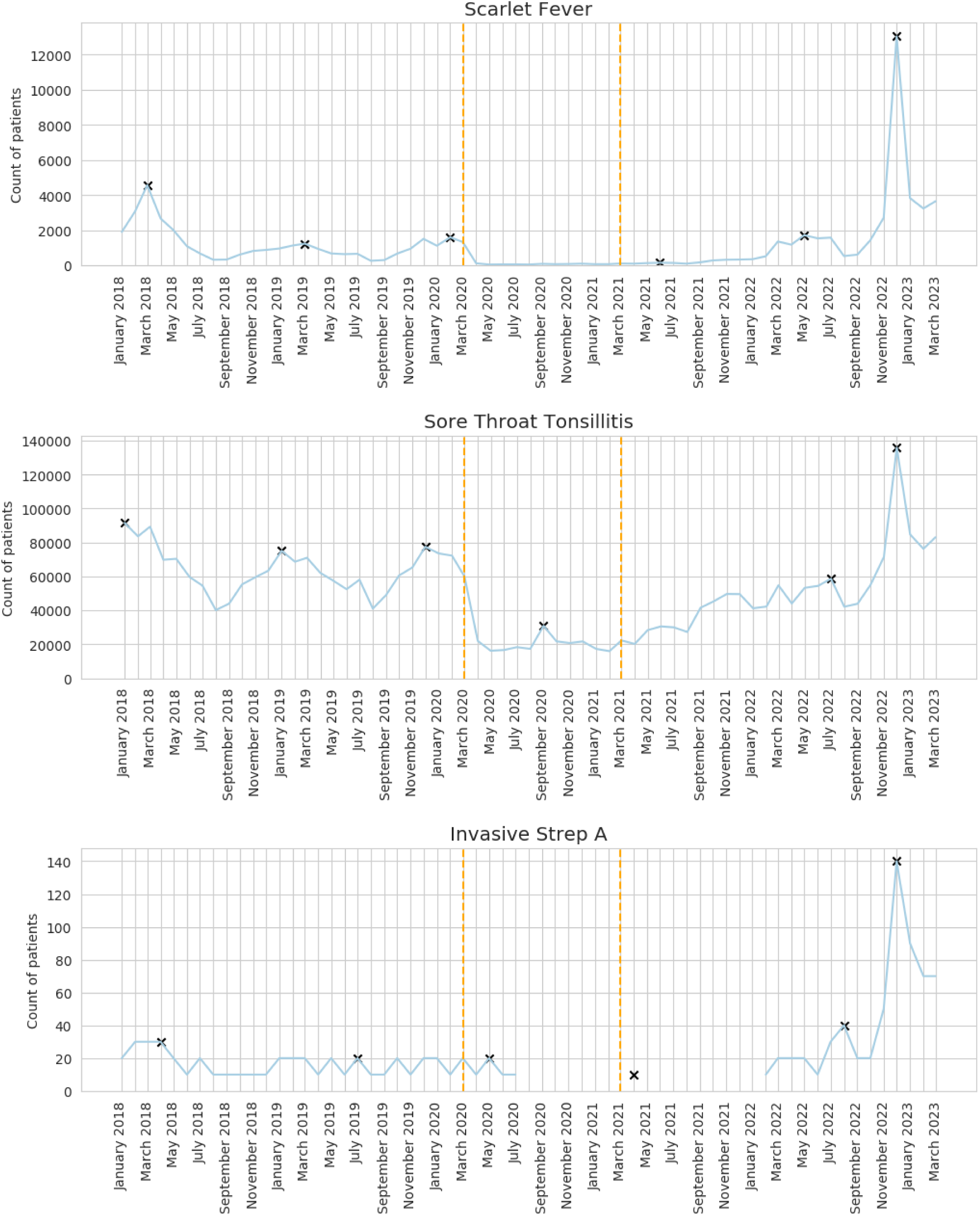
Monthly count of patients with scarlet fever, sore throat/tonsillitis, or iGAS. Vertical lines represent the start of the COVID-19 restriction period (April 2020) and recovery period (April 2021). The maximum value of each season (September through August) is annotated with an x. Gaps indicate the data was redacted due to low counts.

Scarlet fever events similarly followed a seasonal pattern which was not observed during the COVID-19 restriction period. The maximum of the 2020/21 season was 150 cases in June 2021, lower than the pre COVID-19 minimum of 260 in August 2019 (Table S2). Cases then peaked in December 2022 with 13,040 events, a rate ratio of 2.68 (CI:2.59 to 2.77) compared to the pre COVID-19 2017/18 peak of 4,570 in March 2018. 90% of the scarlet fever codes also had an antibiotic prescription up to 7 days prior or 14 days after the clinical event (Table S7).

Recorded iGAS events peaked at 140 in December 2022, a rate ratio of 4.37 (CI:2.94 to 6.48) compared to the 2017/18 peak (Table S2) of 30. Only 29% of scarlet fever codes had an antibiotic prescription 7 days prior or 14 days after the clinical event in the primary care record (Table S7).

Overall, recording of codes for any GAS indication in primary care peaked in December 2022 at 146,260 (5.7 per 1,000). This represented a 1.5-fold increase from the last comparably high season in 2017/18 (93,340 (3.9 per 1,000)) (Table S2).

### Antibiotics used to treat GAS infection

Figure 2 shows counts of antibiotics prescribed with a GAS clinical indication by antibiotic group. The percentage of all prescriptions that had a GAS indication varied by antibiotic, with phenoxymethylpenicillin (Group 1) being the highest in March 2023 at 41%, and flucloxacillin (Group 2) the lowest at less than 1% (Table S7). The same plot of antibiotics prescribed with or without relevant GAS indication (Figure S2), and other metrics are available in the dashboard and the supplemental for context. Prior to COVID-19 restrictions, the highest level of Group 1 prescribing with GAS clinical indication was 48,990 patients in March 2018 followed by seasonal highs of 35,680 patients in January 2019 and 37,910 in December 2019. The 2020/21 season peak of 15,610 in September 2020 was lower than any pre COVID-19 minimum. Counts increased over 2021/22 but without a distinct seasonal pattern. The peak of 71,440 patients in December 2022 compared to the 2017/18 peak had a rate ratio of 1.37 (CI:1.35 to 1.38) (Table S3).

**Figure 2:**
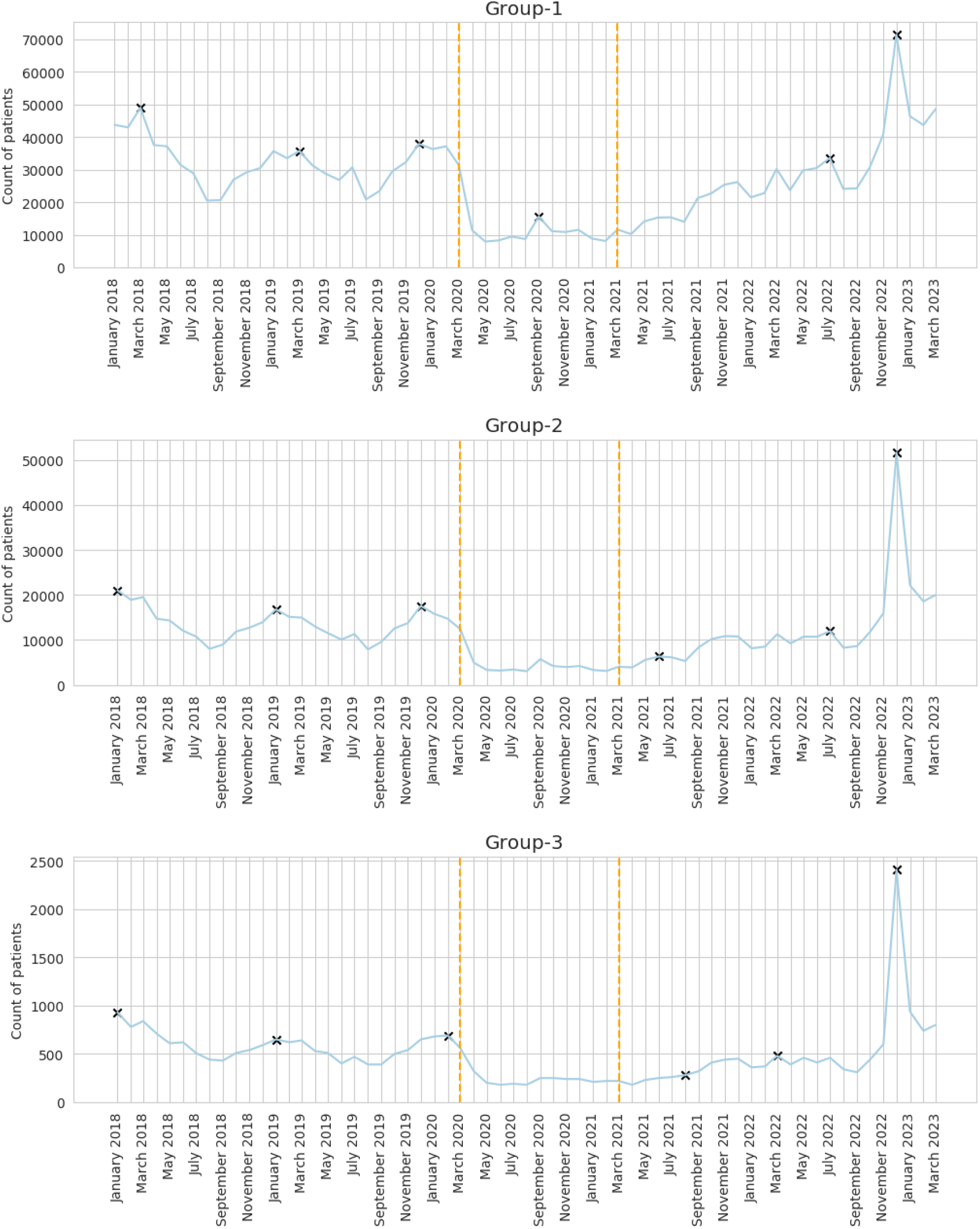
Monthly count of patients with an antibiotic prescription and a record of scarlet fever, sore throat/tonsillitis, or iGAS up to 14 days prior or 7 days after the prescribing event. Antibiotics are grouped into: phenoxymethylpenicillin (Group 1) as the first line antibiotic for GAS infection, macrolides, amoxicillin and flucloxacillin (Group 2) as the antibiotics recommended in case of non-availability or penicillin allergy, and cefalexin and co-amoxiclav (Group 3) as reserved broad-spectrum. Vertical lines represent the start of the COVID-19 restriction period (April 2020) and recovery period (April 2021). The maximum values of each season (September through August) are annotated with an x.

In December 2022, prescribing of Group 2 antibiotics with GAS indication (51,690) was 2.30 (CI:2.26 to 2.34) times the previous high (21,000 in January 2018) and Group 3 antibiotics (2,410) was 2.42 (CI:2.24 to 2.61) times the previous high (930 in January of 2018). Comparing the 2017/18 peak to the 2022 peak for individual antibiotics with a GAS indication (Table S4), azithromycin had the largest increase in rate (RR 7.37 (CI:6.22 to 8.74)), followed by cefalexin (3.81 (CI:3.33 to 4.35)), amoxicillin (2.53 (CI:2.47 to 2.59)), clarithromycin (2.16 (CI:2.10 to 2.22)), co-amoxiclav (1.85 (CI:1.69 to 2.04)), erythromycin (1.71 (CI:1.65 to 1.78)), flucloxacillin (1.49 (CI:1.30 to 1.70)), then phenoxymethylpenicillin (1.37 (CI:1.35 to 1.38)).

### Breakdowns by Demographic Subgroups

The crude rate of phenoxymethylpenicillin prescribed with GAS indication over time (Figure 3) increased with increasing deprivation and was lowest in London, the South West and South East. The rate was highest in those of Mixed ethnicity, followed by South Asian and White, then Black and Other. These patterns were similar throughout the study period.

**Figure 3:**
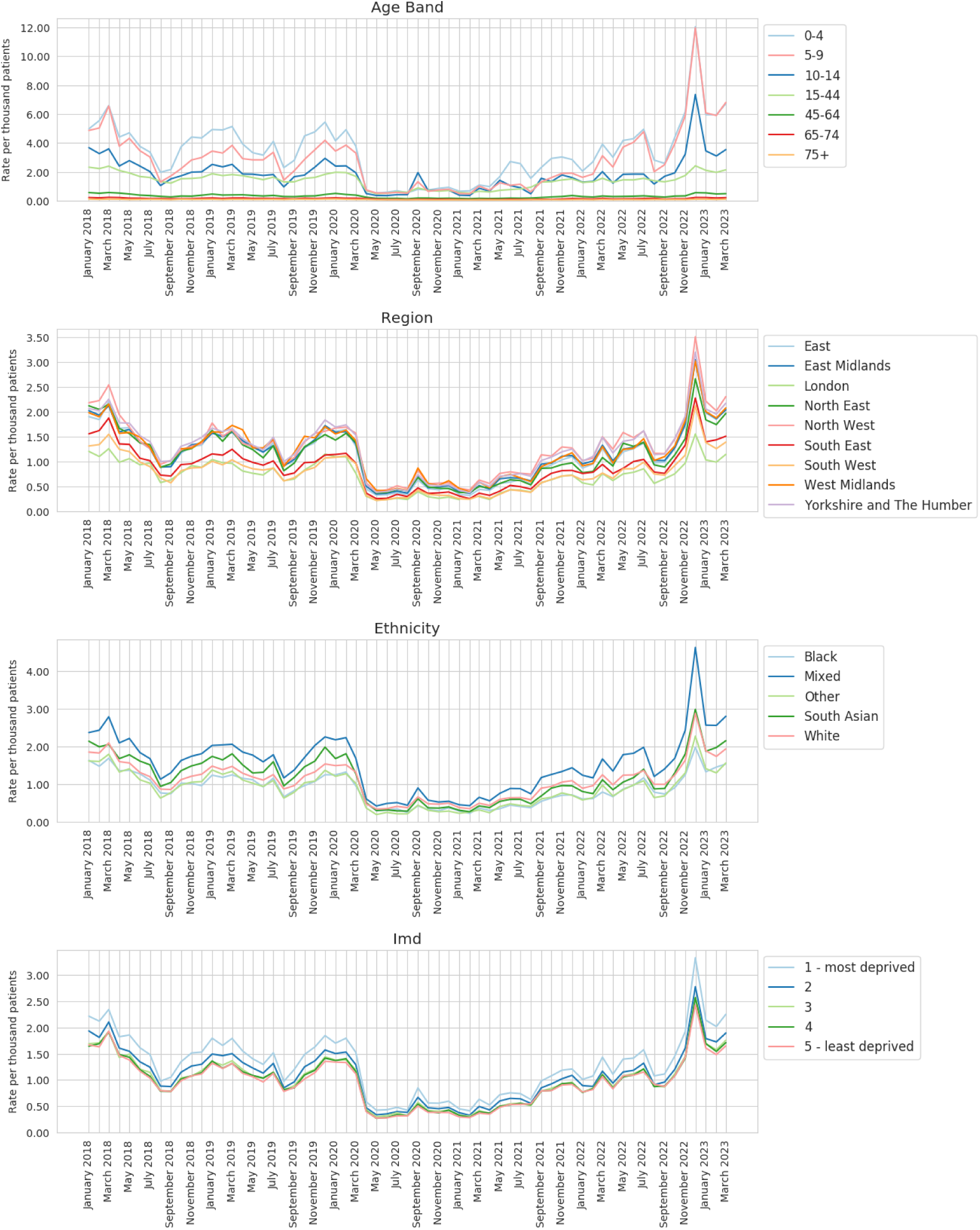
Monthly rate per 1,000 patients with a recorded prescription of phenoxymethylpenicillin and a record of scarlet fever, sore throat/tonsillitis, or iGAS up to 14 days prior or 7 days after the prescribing event by demographic groups

The rate increased with decreasing age, with 0-4 year olds displaying the highest rate throughout the study period. During the 2018/2019 and 2019/20 seasons, the rate of prescribing was higher in 0-4 year olds than 5-9 year olds. During the high seasons (2017/18 and 2022/23) prescribing with GAS indication was similar in the 0-4 and 5-9 age bands (Table S5).

### Dashboard

Our regularly updated dashboards contain more detailed breakdowns for each type of GAS infection and associated antibiotics https://reports.opensafely.org/reports/scarlet-fever-and-invasive-group-a-strep-cases-throughout-the-covid-19-pandemic/, https://reports.opensafely.org/reports/scarlet-fever-and-invasive-group-a-strep-cases-throughout-the-covid-19-pandemic-weekly/. Figure 4 is an example of a more detailed graph, displaying trends over time for the most commonly used sore throat/tonsillitis codes. “Tonsillitis” is the most commonly recorded code over the study period. The individual code trends generally follow the total trend, peaking in winter and dropping during the COVID-19 restriction period. Prior to COVID-19 restrictions, “Pain in throat” made up around 12% of total codes, but spiked to 20% in April 2020. The proportion gradually decreased back to 13% during COVID-19 restrictions, before spiking back to 20% in the December 2022 peak.

**Figure 4:**
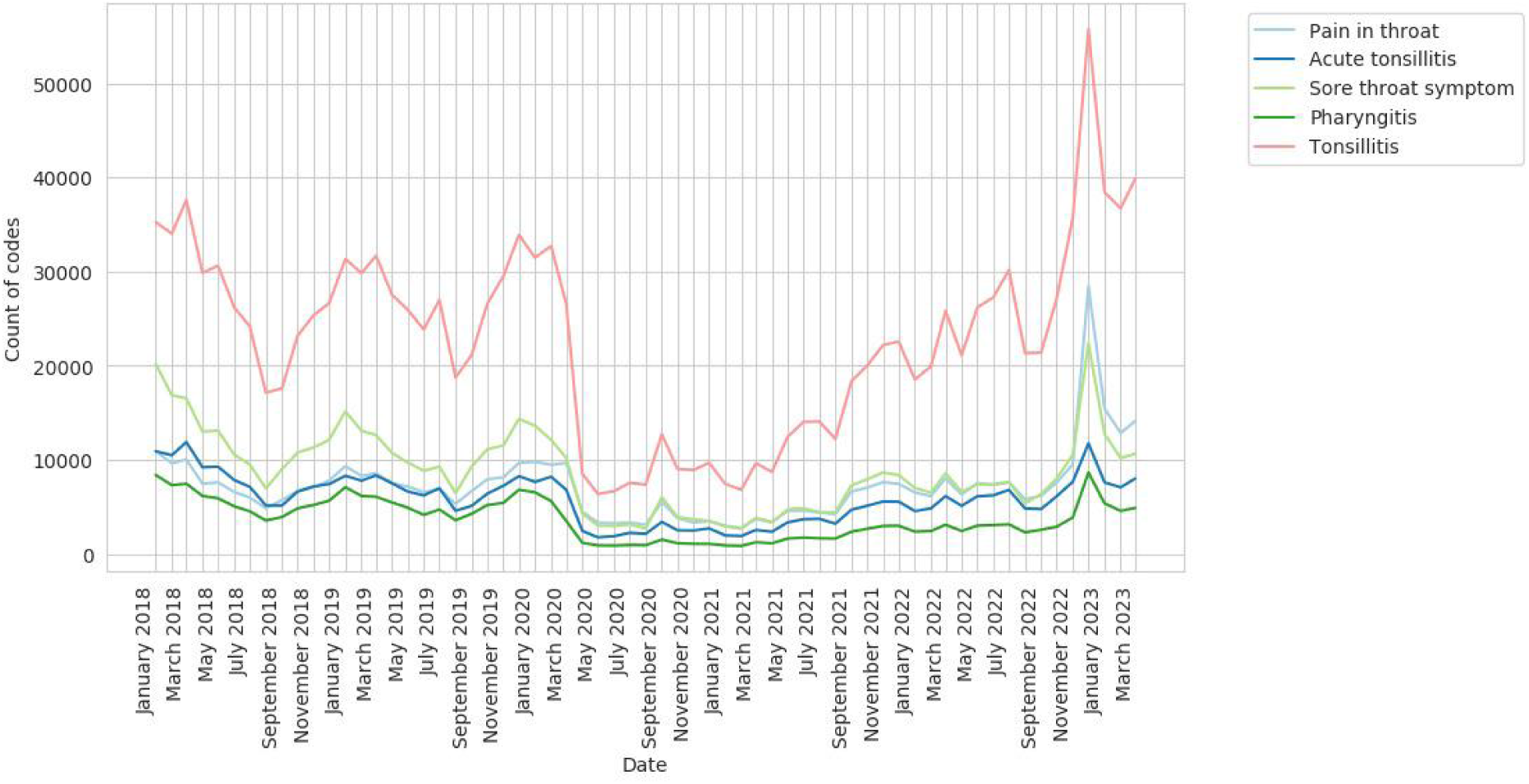
Monthly count of sore throat/tonsillitis code usage

## Discussion

### Statement of principal findings

Recording of codes for GAS indications in primary care peaked in December 2022 at 146,260 (5.7 per 1,000). This represented a 1.5-fold increase from the last comparably high season in 2017/18 (93,340 (3.9 per 1,000)). Prior to COVID-19, GAS events followed a seasonal pattern, peaking sometime between December and March. During the period of COVID-19 restrictions, there was a sharp drop in recording of cases and treatments, with the maximum counts and rates lower than any pre-COVID-19 minimum and no distinct seasonal pattern.

Prescriptions of first line antibiotics in those with a record of a GAS infection peaked at 2.80 per 1,000 (RR 1.37 (CI:1.35 to 1.38)), alternative antibiotics at 2.03 per 1,000 (RR (CI:2.26 to 2.34)), and reserved antibiotics at 0.09 per 1,000 (RR 2.42 (CI:2.24 to 2.61)). For individual antibiotics, azithromycin with GAS indication showed the greatest relative increase (RR 7.37 (CI:6.22 to 8.74)).This followed a sharp drop in recording of cases and associated prescriptions during the period of COVID-19 restrictions where the maximum count and rates were lower than any pre COVID-19 minimum.

### Findings in context

Scarlet fever and iGAS are notifiable diseases in England, meaning that registered practitioners have a statutory duty to report suspected cases to the local council or health protection team by submitting a notification form or by phone [15]. UKHSA publishes weekly and summary notification of infectious disease (NOIDs) reports for England and Wales, with detailed breakdowns by region, county and local authority. From September 12th to December 4th 2022, there were 659 notifications of iGAS in England (1.2 per 100,000) and 6,601 (11.7 per 100,000) notifications of scarlet fever [16]. In OpenSAFELY-TPP from September 1st to November 30th 2022, there were 90 recorded iGAS cases (average rate 0.4 per 100,000) and 4,750 recorded scarlet fever cases (average rate 15 per 100,000). The lower rate of iGAS recording in primary care data is expected as iGAS cases would normally be handled in secondary care. The higher rate of scarlet fever recording is also not unexpected, as disease notifications may be subject to delays in reporting and/or additional adjudication. Our report includes all sore throat/tonsillitis, scarlet fever, and iGAS codelists to provide a sensitive measure more likely to provide an early signal of changes in case counts for decision makers. Further understanding how NOIDS data compares to data in the primary care record is an area for future research.

Daily GAS incidence rates are reported in the GP in-hours syndromic surveillance bulletins, [17] weekly reports published by UKHSA using a sample of around 650 practices covering 7 million registered patients in England. They include breakdowns by age band and UKHSA region, and similar to our OpenSAFELY data, diagnoses may not be laboratory confirmed. Our overall scarlet fever findings are comparable. Our combined clinical indicator (scarlet fever, sore throat/tonsilitis and iGAS) shows a higher rate due to including a broader range of clinical codes/conditions than the most similar indicators in the UKHSA report. Our

OpenSAFELY-TPP report complements the UKHSA reports by: 1) covering a larger sample of practices; 2) including tonsillitis in our sore throat codelist (Figure 4), which may decrease our specificity, but increases our sensitivity; 3) linking antibiotic prescribing codes to diagnosis codes at the patient level, allowing us to monitor GAS related antibiotic prescribing; 4) providing more detailed breakdowns (whereas the GP in-hours report provides a quick summary for GPs of which syndromes might be above or below baseline); and 5) sharing all our analytic code and codelists openly for examination, comparison and re-use (Table S1).

Lower levels of circulating viruses during the COVID-19 restrictions may have contributed to a spike in GAS cases in December 2022. Although two years have passed since restrictions began to ease in England, the ongoing monitoring of infectious diseases and the prescription of associated treatments will continue to be important while non-seasonal patterns of viruses are still observed.

### Strengths and weaknesses

The main strength of this study is the speed and scale of its delivery to the NHS outbreak team. The time from project approval to first report was 7 days. This study was implemented across the full EHR coded data covering 40% of GP practices in England. The report was, and can in future, be delivered on a weekly basis, providing a near real-time warning system in the case of future outbreaks and pressures on antibiotic supply. As this dashboard was developed for a small outbreak team, user research into how clinicians and decision makers could best make use of this or similar surveillance reports could improve future versions.

In the absence of a clear indicator of group A strep throat, a codelist for sore throat/tonsillitis was developed where an alternative cause is not stated (e.g.Staphylococcal tonsillitis). As these symptoms can still often be caused by viruses, including COVID-19, we used the clinical codelists in combination with codelists for antibiotics recommended for treatment of GAS to improve specificity. In March 2023, 75% of these codes were associated with an antibiotic prescription, but cases may still be viral and not bacterial. It is likely that our codelists capture some non-GAS events, which is why in addition to reporting the total number of possible GAS events, we split the clinical events into iGAS, scarlet fever, and sore throat/tonsillitis. Additionally, our approach also relies on a clinician adding an appropriate clinical code to indicate a diagnosis, but coding of consultations varies, and some consultations may not be coded.

Our study looks at antibiotic prescriptions issued, however prescriptions recorded in the primary care record may not always be dispensed or in some cases the dispensed item may differ from the prescribed item due to the use of a Serious Shortage Protocol [9]. SSPs for phenoxymethylpenicillin were in place in the UK from December 15th 2022 until May 12th 2023, allowing for other formulations of phenoxymethylpenicillin, or substitution with amoxicillin, flucloxacillin, cefalexin, co-amoxiclav, erythromycin and, up until mid January, azithromycin and clarithromycin. In two recent Freedom of Information requests [18,19], we found that from December 2022 through March 2023, 35,458 items were dispensed under these SSPs across England, 23,583 of which were not a phenoxymethylpenicillin formulation (Table S8) https://github.com/opensafely/strepa_scarlet/tree/main/analysis/ssp-analysis. In those same months in OpenSAFELY-TPP (40% of GP practices in England), there was a record of over 530,000 phenoxymethylpenicillin prescriptions. If we assume OpenSAFELY-TPP prescribing is representative of overall prescribing, then the substitution of phenoxymethylpenicillin with a non-phenoxymethylpenicillin alternative would represent less than 2% of phenoxymethylpenicillin prescribing. Therefore, although this analysis is based on prescriptions and not dispensings, SSPs likely had minimal impact on the findings. There is currently no national data linking prescriptions to dispensed items [20]; we encourage collection of this data in future to aid the understanding of the impact of SSPs in future research.

In this study we did not identify infections through diagnostic tests, test results or clinical scoring tools. Throat swabs are recommended where there is diagnostic uncertainty [7], but diagnosis of scarlet fever and other GAS infections remains based on clinical criteria. Clinical scoring tools such as feverPAIN are recommended for determining the likelihood antibiotics would help a patient, and during the December 2022 outbreak, the feverPAIN prescribing threshold was lowered from 4 to 3 [7]. Future research could investigate the availability and usefulness of relevant diagnostic tests and clinical scoring tools in electronic health records, and whether they can help improve the specificity of our analyses. Future work could also make use of additional data sources such as onward presentations at hospitals.

### Policy implications

OpenSAFELY is a secure health analytics platform that allows near real-time analysis of pseudonymised primary care patient records in England to support response to the COVID-19 pandemic. We have previously shown that the OpenSAFELY platform can be used for rapid audit and feedback [10]. Here we have demonstrated that OpenSAFELY can support the response to an outbreak of infectious disease associated with the pandemic, giving detailed information on disease recording and prescribing in general practice. This software framework can be reused or repurposed to provide near real-time surveillance for future disease outbreaks or prescribing of any medication.

### Conclusion

We found that prior to COVID-19, sore throat/tonsillitis, scarlet fever, and iGAS events followed a seasonal pattern, peaking sometime between December and March. During the period of COVID-19 restrictions, there was a sharp drop, with the maximum counts and rates lower than any pre COVID-19 minimum. Cases increased over 2021/22 but without a distinct seasonal pattern, until peaking in December 2022. Primary care data can supplement existing infectious disease surveillance reports by linking with patient level prescribing data. We produced a live updating dashboard with more detailed breakdowns and sensitive codelists to provide greater context to relevant analysts and outbreak teams.

## Supporting information

strepa_supplemental

## Administrative

## Acknowledgements

We are very grateful for all the support received from the TPP Technical Operations team throughout this work, and for generous assistance from the information governance and database teams at NHS England and the NHS England Transformation Directorate

## Information governance and ethical approval

NHS England is the data controller for OpenSAFELY-TPP; TPP is the data processor; all study authors using OpenSAFELY have the approval of NHS England [21]. This implementation of OpenSAFELY is hosted within the TPP environment which is accredited to the ISO 27001 information security standard and is NHS IG Toolkit compliant [22].

Patient data has been pseudonymised for analysis and linkage using industry standard cryptographic hashing techniques; all pseudonymised datasets transmitted for linkage onto OpenSAFELY are encrypted; access to the platform is via a virtual private network (VPN) connection, restricted to a small group of researchers; the researchers hold contracts with NHS England and only access the platform to initiate database queries and statistical models; all database activity is logged; only aggregate statistical outputs leave the platform environment following best practice for anonymisation of results such as statistical disclosure control for low cell counts [23].

The service adheres to the obligations of the UK General Data Protection Regulation (UK GDPR) and the Data Protection Act 2018. The service previously operated under notices initially issued in February 2020 by the the Secretary of State under Regulation 3(4) of the Health Service (Control of Patient Information) Regulations 2002 (COPI Regulations), which required organisations to process confidential patient information for COVID-19 purposes; this set aside the requirement for patient consent [24]. As of 1 July 2023, the Secretary of State has requested that NHS England continue to operate the Service under the COVID-19 Directions 2020 [25]. In some cases of data sharing, the common law duty of confidence is met using, for example, patient consent or support from the Health Research Authority Confidentiality Advisory Group [26].

Taken together, these provide the legal bases to link patient datasets on the OpenSAFELY platform. GP practices, from which the primary care data are obtained, are required to share relevant health information to support the public health response to the pandemic, and have been informed of the OpenSAFELY analytics platform.

This service evaluation study was supported by Dr. Kieran Hand, National Pharmacy & Prescribing Clinical Lead for antimicrobial resistance at NHS England, as senior sponsor. This study was approved by the Health Research Authority (REC reference 20/LO/0651).

## Data sharing

All data were linked, stored and analysed securely within the OpenSAFELY platform https://opensafely.org/. Data include pseudonymized data such as coded diagnoses, medications and physiological parameters. No free text data are included. All code is shared openly for review and re-use under MIT open license https://github.com/opensafely/strepa_scarlet. Detailed pseudonymised patient data is potentially re-identifiable and therefore not shared. We rapidly delivered the OpenSAFELY data analysis platform without prior funding to deliver timely analyses on urgent research questions in the context of the global Covid-19 health emergency: now that the platform is established we are developing a formal process for external users to request access in collaboration with NHS England; details of this process are available at https://www.opensafely.org/onboarding-new-users/.

## Guarantor

BM is guarantor.

## Contributorship

**Conceptualization:** C.C., L.F., C.W., R.C., E.B., K.H., and B.M.

**Data curation:** C.C., L.F., B.F.B.-C., D.E., P.I., I.D., S.C.B., S.D., T.W., G.H., L.B., T.O.D., S.M., R.M.S., A.M., C.B., J.C., J.P., F.H., and S.H.

**Formal analysis:** C.C., L.F., and C.W.

**Funding acquisition:** B.G.

**Methodology:** C.C., L.F., C.W., V.S., A.D.B., R.C., and B.M.

**Project administration:** A.M. and L.C.H.

**Resources:** B.F.B.-C., D.E., P.I., I.D., S.C.B., S.D., T.W., G.H., L.B., T.O.D., S.M., R.M.S., A.M., C.B., J.C., J.P., F.H., and S.H.

**Software:** B.F.B.-C., D.E., P.I., I.D., S.C.B., S.D., T.W., G.H., L.B., T.O.D., S.M., R.M.S., C.B., J.C., J.P., F.H., and S.H.

**Supervision:** H.J.C., R.H., and B.M.

**Validation:** C.C., L.F., and C.W.

**Visualization:** C.C. and L.F.

**Writing - original draft:** C.C., L.F., and C.W.

**Writing - review & editing:** C.C., L.F., C.W., H.J.C., E.B., A.M., and B.M.

