## Supplementary material for "Group A streptococcal cases and treatments during the COVID-19 pandemic and 2022 outbreak: a retrospective cohort study in England using OpenSAFELY-TPP": strepa_supplemental

**Figure S1.** Population denominator in OpenSAFELY-TPP from January 2018 through March 2023. Patients who have died, or have missing age or sex have been excluded

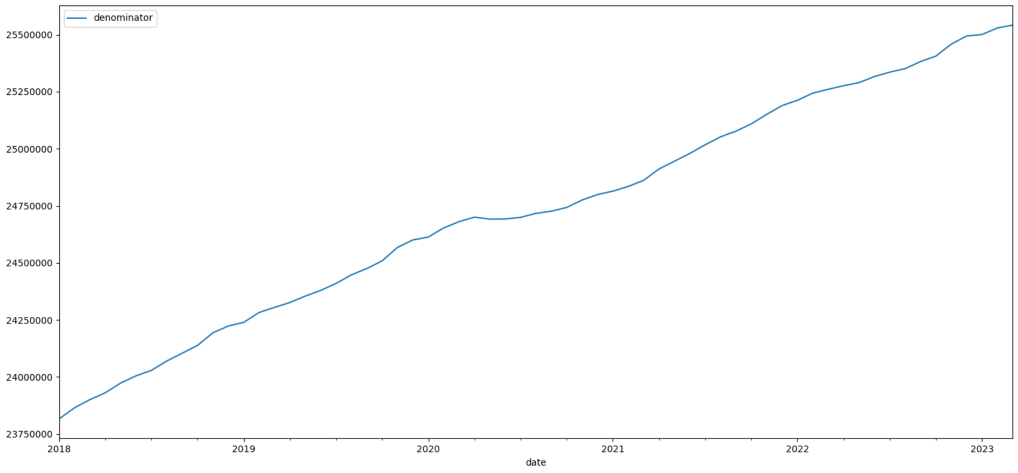


**Table S1.** Codelists used in this study

| Codelist type | Codelist description | Link |
|---|---|---|
| Antibiotic | Phenoxymethylpenicillin | https://codelists.opensafely.org/codelist/opensafely/phenoxymethylpenicillin-oral-preparations-only/14b427f8/ |
| | Amoxicillin | https://codelists.opensafely.org/codelist/opensafely/amoxicillin-oral/164af813/ |
| | Clarithromycin | https://codelists.opensafely.org/codelist/opensafely/clarithromycin-oral/080684b6/ |
| | Erythromycin | https://codelists.opensafely.org/codelist/opensafely/erythromycin-oral/7d3b84dd/ |
| | Azithromycin | https://codelists.opensafely.org/codelist/opensafely/azithromycin-oral/42faf594/ |
| | Flucloxacillin | https://codelists.opensafely.org/codelist/opensafely/flucloxacillin-oral/35dbdb0f/ |
| | Cefalexin | https://codelists.opensafely.org/codelist/opensafely/cefalexin-oral/4eeb4e44/ |
| | Co-amoxiclav | https://codelists.opensafely.org/codelist/opensafely/co-amoxiclav-oral/67fac179/ |
| Clinical event | Scarlet Fever | https://opencodelists.org/codelist/opensafely/scarlet-fever/1ba70f02/ |
| | Sore throat/tonsillitis | https://opencodelists.org/codelist/opensafely/group-a-streptococcal-sore-throat/2924ced0/ |
| | Invasive Strep A | https://.opencodelists.org/codelist/opensafely/invasive-group-a-strep/42344205/ |

**Table S2:** The max and min count and rate per 1,000 patients for scarlet fever, sore throat/tonsillitis, or iGAS in each season (September through August). Counts have been rounded to the nearest 10 and the rate computed with the rounded number.

| | gas_year | 2017/18 | | 2018/19 | | 2019/20 | | 2020/21 | | 2021/22 | | 2022/23 | | 2022/23 v 2017/18 |
|---|---|---|---|---|---|---|---|---|---|---|---|---|---|---|
| | | Count | Rate (95% CI) | Count | Rate (95% CI) | Count | Rate (95% CI) | Count | Rate (95% CI) | Count | Rate (95% CI) | Count | Rate (95% CI) | Rate Ratio (95% CI) |
| name | type | | | | | | | | | | | | | |
| Invasive Strep A | max | 30 | 0.00 (0.00 to 0.00) | 20 | 0.00 (0.00 to 0.00) | 20 | 0.00 (0.00 to 0.00) | 10 | 0.00 (0.00 to 0.00) | 40 | 0.00 (0.00 to 0.00) | 140 | 0.01 (0.00 to 0.01) | 4.37 (2.94 to 6.48) |
| | min | 10 | 0.00 (0.00 to 0.00) | 10 | 0.00 (0.00 to 0.00) | 10 | 0.00 (0.00 to 0.00) | 10 | 0.00 (0.00 to 0.00) | 10 | 0.00 (0.00 to 0.00) | 20 | 0.00 (0.00 to 0.00) | 1.89 (0.89 to 4.05) |
| Scarlet Fever | max | 4570 | 0.19 (0.19 to 0.20) | 1240 | 0.05 (0.05 to 0.05) | 1600 | 0.06 (0.06 to 0.07) | 150 | 0.01 (0.01 to 0.01) | 1720 | 0.07 (0.06 to 0.07) | 13040 | 0.51 (0.50 to 0.52) | 2.68 (2.59 to 2.77) |
| | min | 320 | 0.01 (0.01 to 0.01) | 260 | 0.01 (0.01 to 0.01) | 50 | 0.00 (0.00 to 0.00) | 70 | 0.00 (0.00 to 0.00) | 170 | 0.01 (0.01 to 0.01) | 610 | 0.02 (0.02 to 0.03) | 1.81 (1.58 to 2.07) |
| Sore Throat Tonsillitis | max | 91510 | 3.84 (3.82 to 3.87) | 75070 | 3.10 (3.07 to 3.12) | 77470 | 3.15 (3.13 to 3.17) | 30860 | 1.25 (1.23 to 1.26) | 58630 | 2.31 (2.30 to 2.33) | 135860 | 5.33 (5.30 to 5.36) | 1.39 (1.38 to 1.40) |
| | min | 40130 | 1.67 (1.65 to 1.68) | 41000 | 1.68 (1.66 to 1.69) | 16170 | 0.65 (0.64 to 0.66) | 15940 | 0.64 (0.63 to 0.65) | 41210 | 1.63 (1.62 to 1.65) | 43880 | 1.73 (1.71 to 1.74) | 1.04 (1.02 to 1.05) |
| Any GAS Clinical Event | max | 93,340 | 3.91 (3.88 to 3.93) | 75,930 | 3.13 (3.11 to 3.15) | 78,830 | 3.20 (3.18 to 3.23) | 30,940 | 1.25 (1.24 to 1.27) | 60,040 | 2.37 (2.35 to 2.39) | 146,260 | 5.74 (5.71 to 5.77) | 1.47 (1.46 to 1.48) |
| | min | 40,420 | 1.68 (1.66 to 1.70) | 41,230 | 1.69 (1.67 to 1.70) | 16,240 | 0.66 (0.65 to 0.67) | 16,000 | 0.64 (0.63 to 0.65) | 41,520 | 1.65 (1.63 to 1.66) | 44,430 | 1.75 (1.73 to 1.77) | 1.04 (1.03 to 1.06) |

**Table S3.** The max and min count and rate per 1,000 patients with an antibiotic prescription and a record of scarlet fever, sore throat/tonsillitis, or iGAS up to 14 days prior or 7 days after the prescribing event for each season (September through August). Antibiotics are grouped as phenoxymethylpenicillin (Group 1) as the first line antibiotic for GAS infection, macrolides, amoxicillin and flucloxacillin (Group 2) as the antibiotics recommended in case of non-availability or penicillin allergy, and cefalexin and co-amoxiclav (Group 3) as reserved broad-spectrum. Counts have been rounded to the nearest 10 and the rate computed with the rounded number.

| | gas_year | 2017/18 | | 2018/19 | | 2019/20 | | 2020/21 | | 2021/22 | | 2022/23 | | 2022/23 v 2017/18 |
|---|---|---|---|---|---|---|---|---|---|---|---|---|---|---|
| | | Count | Rate (95% CI) | Count | Rate (95% CI) | Count | Rate (95% CI) | Count | Rate (95% CI) | Count | Rate (95% CI) | Count | Rate (95% CI) | Rate Ratio (95% CI) |
| name | type | | | | | | | | | | | | | |
| Group-1 | max | 48990 | 2.05 (2.03 to 2.07) | 35680 | 1.47 (1.46 to 1.49) | 37910 | 1.54 (1.53 to 1.56) | 15610 | 0.63 (0.62 to 0.64) | 33560 | 1.32 (1.31 to 1.34) | 71440 | 2.80 (2.78 to 2.82) | 1.37 (1.35 to 1.38) |
| | min | 20510 | 0.85 (0.84 to 0.86) | 20830 | 0.85 (0.84 to 0.86) | 7950 | 0.32 (0.31 to 0.33) | 8140 | 0.33 (0.32 to 0.33) | 21280 | 0.85 (0.84 to 0.86) | 24290 | 0.96 (0.94 to 0.97) | 1.12 (1.10 to 1.14) |
| Group-2 | max | 21000 | 0.88 (0.87 to 0.89) | 16790 | 0.69 (0.68 to 0.70) | 17560 | 0.71 (0.70 to 0.72) | 6300 | 0.25 (0.25 to 0.26) | 11950 | 0.47 (0.46 to 0.48) | 51690 | 2.03 (2.01 to 2.04) | 2.30 (2.26 to 2.34) |
| | min | 7990 | 0.33 (0.32 to 0.34) | 7860 | 0.32 (0.31 to 0.33) | 3010 | 0.12 (0.12 to 0.13) | 3060 | 0.12 (0.12 to 0.13) | 8120 | 0.32 (0.32 to 0.33) | 8590 | 0.34 (0.33 to 0.35) | 1.02 (0.99 to 1.05) |
| Group-3 | max | 930 | 0.04 (0.04 to 0.04) | 650 | 0.03 (0.02 to 0.03) | 690 | 0.03 (0.03 to 0.03) | 280 | 0.01 (0.01 to 0.01) | 480 | 0.02 (0.02 to 0.02) | 2410 | 0.09 (0.09 to 0.10) | 2.42 (2.24 to 2.61) |
| | min | 440 | 0.02 (0.02 to 0.02) | 390 | 0.02 (0.01 to 0.02) | 180 | 0.01 (0.01 to 0.01) | 180 | 0.01 (0.01 to 0.01) | 320 | 0.01 (0.01 to 0.01) | 310 | 0.01 (0.01 to 0.01) | 0.67 (0.58 to 0.77) |

**Figure S2:** Monthly count of patients with an antibiotic prescription (with or without GAS indication). Antibiotics have been grouped into Group 1, Group 2, and Group 3. Vertical lines represent the start of the COVID-19 restriction period (April 2020) and recovery period (April 2021). The maximum value of each season (September through August) is annotated with an x.

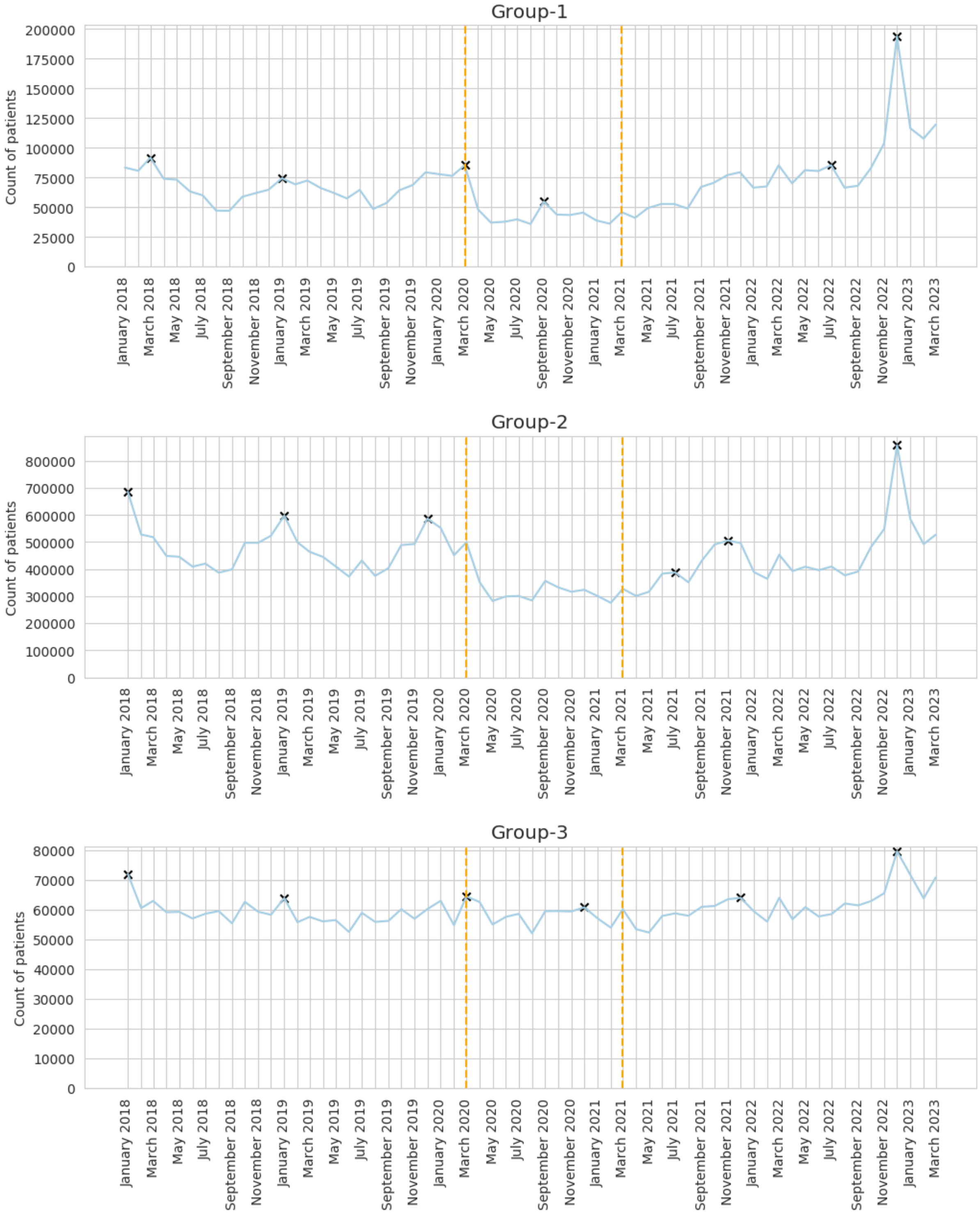

**Table S4.** The max and min count and rate per 1,000 patients with an antibiotic prescription and a record of scarlet fever, sore throat/tonsillitis, or iGAS up to 14 days prior or 7 days after the prescribing event for each season (September through August). Counts have been rounded to the nearest 10 and the rate computed with the rounded number.

| | gas_year | 2017/18 | | 2018/19 | | 2019/20 | | 2020/21 | | 2021/22 | | 2022/23 | | 2022/23 v 2017/18 |
|---|---|---|---|---|---|---|---|---|---|---|---|---|---|---|
| | | Count | Rate (95% CI) | Count | Rate (95% CI) | Count | Rate (95% CI) | Count | Rate (95% CI) | Count | Rate (95% CI) | Count | Rate (95% CI) | Rate Ratio (95% CI) |
| name | type | | | | | | | | | | | | | |
| Amoxicillin | max | 9880 | 0.41 (0.41 to 0.42) | 7940 | 0.33 (0.32 to 0.33) | 8440 | 0.34 (0.34 to 0.35) | 2660 | 0.11 (0.10 to 0.11) | 4630 | 0.18 (0.18 to 0.19) | 26770 | 1.05 (1.04 to 1.06) | 2.53 (2.47 to 2.59) |
| | min | 3120 | 0.13 (0.13 to 0.13) | 3020 | 0.12 (0.12 to 0.13) | 920 | 0.04 (0.03 to 0.04) | 1030 | 0.04 (0.04 to 0.04) | 2820 | 0.11 (0.11 to 0.12) | 3370 | 0.13 (0.13 to 0.14) | 1.02 (0.98 to 1.08) |
| Azithromycin | max | 150 | 0.01 (0.01 to 0.01) | 130 | 0.01 (0.00 to 0.01) | 120 | 0.00 (0.00 to 0.01) | 50 | 0.00 (0.00 to 0.00) | 100 | 0.00 (0.00 to 0.00) | 1180 | 0.05 (0.04 to 0.05) | 7.37 (6.22 to 8.74) |
| | min | 60 | 0.00 (0.00 to 0.00) | 60 | 0.00 (0.00 to 0.00) | 30 | 0.00 (0.00 to 0.00) | 20 | 0.00 (0.00 to 0.00) | 40 | 0.00 (0.00 to 0.00) | 80 | 0.00 (0.00 to 0.00) | 1.26 (0.90 to 1.77) |
| Cefalexin | max | 270 | 0.01 (0.01 to 0.01) | 190 | 0.01 (0.01 to 0.01) | 210 | 0.01 (0.01 to 0.01) | 90 | 0.00 (0.00 to 0.00) | 160 | 0.01 (0.01 to 0.01) | 1100 | 0.04 (0.04 to 0.05) | 3.81 (3.33 to 4.35) |
| | min | 120 | 0.00 (0.00 to 0.01) | 100 | 0.00 (0.00 to 0.00) | 40 | 0.00 (0.00 to 0.00) | 60 | 0.00 (0.00 to 0.00) | 100 | 0.00 (0.00 to 0.00) | 90 | 0.00 (0.00 to 0.00) | 0.71 (0.54 to 0.93) |
| Clarithromycin | max | 7120 | 0.30 (0.29 to 0.31) | 6110 | 0.25 (0.25 to 0.26) | 6450 | 0.26 (0.26 to 0.27) | 2680 | 0.11 (0.10 to 0.11) | 5530 | 0.22 (0.21 to 0.22) | 16460 | 0.65 (0.64 to 0.66) | 2.16 (2.10 to 2.22) |
| | min | 3300 | 0.14 (0.13 to 0.14) | 3410 | 0.14 (0.14 to 0.15) | 1580 | 0.06 (0.06 to 0.07) | 1490 | 0.06 (0.06 to 0.06) | 3730 | 0.15 (0.14 to 0.15) | 4110 | 0.16 (0.16 to 0.17) | 1.18 (1.13 to 1.24) |
| Co Amoxiclav | max | 660 | 0.03 (0.03 to 0.03) | 480 | 0.02 (0.02 to 0.02) | 500 | 0.02 (0.02 to 0.02) | 200 | 0.01 (0.01 to 0.01) | 330 | 0.01 (0.01 to 0.01) | 1310 | 0.05 (0.05 to 0.05) | 1.85 (1.69 to 2.04) |
| | min | 310 | 0.01 (0.01 to 0.01) | 280 | 0.01 (0.01 to 0.01) | 120 | 0.00 (0.00 to 0.01) | 120 | 0.00 (0.00 to 0.01) | 220 | 0.01 (0.01 to 0.01) | 220 | 0.01 (0.01 to 0.01) | 0.67 (0.57 to 0.80) |
| Erythromycin | max | 3690 | 0.15 (0.15 to 0.16) | 2370 | 0.10 (0.09 to 0.10) | 2290 | 0.09 (0.09 to 0.10) | 870 | 0.03 (0.03 to 0.04) | 1540 | 0.06 (0.06 to 0.06) | 6740 | 0.26 (0.26 to 0.27) | 1.71 (1.65 to 1.78) |
| | min | 1280 | 0.05 (0.05 to 0.06) | 1110 | 0.05 (0.04 to 0.05) | 380 | 0.02 (0.01 to 0.02) | 420 | 0.02 (0.02 to 0.02) | 880 | 0.03 (0.03 to 0.04) | 850 | 0.03 (0.03 to 0.04) | 0.63 (0.58 to 0.69) |
| Flucloxacillin | max | 340 | 0.01 (0.01 to 0.02) | 290 | 0.01 (0.01 to 0.01) | 310 | 0.01 (0.01 to 0.01) | 140 | 0.01 (0.00 to 0.01) | 250 | 0.01 (0.01 to 0.01) | 540 | 0.02 (0.02 to 0.02) | 1.49 (1.30 to 1.70) |
| | min | 230 | 0.01 (0.01 to 0.01) | 190 | 0.01 (0.01 to 0.01) | 90 | 0.00 (0.00 to 0.00) | 70 | 0.00 (0.00 to 0.00) | 130 | 0.01 (0.00 to 0.01) | 180 | 0.01 (0.01 to 0.01) | 0.74 (0.61 to 0.90) |
| Phenoxymethyl penicillin | max | 48990 | 2.05 (2.03 to 2.07) | 35680 | 1.47 (1.46 to 1.49) | 37910 | 1.54 (1.53 to 1.56) | 15610 | 0.63 (0.62 to 0.64) | 33560 | 1.32 (1.31 to 1.34) | 71440 | 2.80 (2.78 to 2.82) | 1.37 (1.35 to 1.38) |
| | min | 20510 | 0.85 (0.84 to 0.86) | 20830 | 0.85 (0.84 to 0.86) | 7950 | 0.32 (0.31 to 0.33) | 8140 | 0.33 (0.32 to 0.33) | 21280 | 0.85 (0.84 to 0.86) | 24290 | 0.96 (0.94 to 0.97) | 1.12 (1.10 to 1.14) |

**Table S5.** The max and min count and rate per 1,000 patients with a phenoxymethylpenicillin prescription and a record of scarlet fever, sore throat/tonsillitis, or iGAS up to 14 days prior or 7 days after the prescribing event for each season (September through August), by age band. Counts have been rounded to the nearest 10 and the rate computed with the rounded number.

| | gas_year | 2017/18 | | 2018/19 | | 2019/20 | | 2021/21 | | 2021/22 | | 2022/23 | | 2022/23 v 2017/18 |
|---|---|---|---|---|---|---|---|---|---|---|---|---|---|---|
| | | Count | Rate (95% CI) | Count | Rate (95% CI) | Count | Rate (95% CI) | Count | Rate (95% CI) | Count | Rate (95% CI) | Count | Rate (95% CI) | Rate Ratio (95% CI) |
| group | type | | | | | | | | | | | | | |
| 0-4 | max | 8530 | 6.59 (6.45 to 6.73) | 6610 | 5.14 (5.01 to 5.26) | 6930 | 5.44 (5.32 to 5.57) | 3310 | 2.71 (2.62 to 2.80) | 5970 | 4.95 (4.83 to 5.08) | 14490 | 12.05 (11.86 to 12.25) | 1.83 (1.78 to 1.88) |
| | min | 2540 | 1.97 (1.89 to 2.05) | 2780 | 2.16 (2.08 to 2.24) | 670 | 0.53 (0.49 to 0.57) | 810 | 0.66 (0.61 to 0.70) | 2490 | 2.05 (1.97 to 2.13) | 3100 | 2.58 (2.49 to 2.67) | 1.31 (1.24 to 1.38) |
| 5-9 | max | 9490 | 6.56 (6.43 to 6.69) | 5580 | 3.83 (3.73 to 3.93) | 6100 | 4.18 (4.08 to 4.28) | 1900 | 1.31 (1.25 to 1.37) | 6860 | 4.78 (4.66 to 4.89) | 17180 | 11.97 (11.79 to 12.14) | 1.82 (1.78 to 1.87) |
| | min | 1880 | 1.30 (1.24 to 1.36) | 2070 | 1.42 (1.36 to 1.48) | 750 | 0.52 (0.48 to 0.55) | 680 | 0.47 (0.43 to 0.51) | 1880 | 1.30 (1.24 to 1.36) | 3580 | 2.49 (2.41 to 2.58) | 1.92 (1.82 to 2.03) |
| 10-14 | max | 4940 | 3.67 (3.57 to 3.78) | 3520 | 2.52 (2.43 to 2.60) | 4200 | 2.93 (2.85 to 3.02) | 2830 | 1.95 (1.87 to 2.02) | 3040 | 2.01 (1.94 to 2.08) | 11300 | 7.36 (7.22 to 7.49) | 2.00 (1.94 to 2.07) |
| | min | 1440 | 1.05 (0.99 to 1.10) | 1350 | 0.95 (0.90 to 1.00) | 510 | 0.35 (0.32 to 0.38) | 530 | 0.36 (0.33 to 0.39) | 1760 | 1.16 (1.10 to 1.21) | 2580 | 1.69 (1.62 to 1.76) | 1.61 (1.51 to 1.72) |
| 15-44 | max | 21780 | 2.39 (2.36 to 2.42) | 17290 | 1.87 (1.84 to 1.90) | 18580 | 1.97 (1.94 to 2.00) | 9010 | 0.93 (0.91 to 0.95) | 15250 | 1.55 (1.53 to 1.58) | 24040 | 2.42 (2.38 to 2.45) | 1.01 (0.99 to 1.03) |
| | min | 12220 | 1.33 (1.31 to 1.35) | 11190 | 1.22 (1.19 to 1.24) | 4550 | 0.48 (0.47 to 0.49) | 4960 | 0.52 (0.51 to 0.53) | 12400 | 1.27 (1.25 to 1.29) | 12950 | 1.31 (1.29 to 1.33) | 0.98 (0.96 to 1.01) |
| 45-64 | max | 3540 | 0.57 (0.55 to 0.58) | 2880 | 0.45 (0.44 to 0.47) | 3220 | 0.50 (0.48 to 0.52) | 1170 | 0.18 (0.17 to 0.19) | 2310 | 0.35 (0.34 to 0.37) | 3640 | 0.56 (0.54 to 0.57) | 0.98 (0.94 to 1.03) |
| | min | 1880 | 0.30 (0.29 to 0.31) | 1630 | 0.26 (0.25 to 0.27) | 830 | 0.13 (0.12 to 0.14) | 870 | 0.13 (0.13 to 0.14) | 1440 | 0.22 (0.21 to 0.23) | 1630 | 0.25 (0.24 to 0.26) | 0.83 (0.78 to 0.89) |
| 65-74 | max | 560 | 0.23 (0.21 to 0.25) | 460 | 0.19 (0.17 to 0.20) | 500 | 0.20 (0.18 to 0.22) | 240 | 0.10 (0.08 to 0.11) | 390 | 0.16 (0.14 to 0.17) | 550 | 0.22 (0.20 to 0.24) | 0.97 (0.86 to 1.09) |
| | min | 340 | 0.14 (0.12 to 0.15) | 320 | 0.13 (0.12 to 0.14) | 170 | 0.07 (0.06 to 0.08) | 160 | 0.06 (0.05 to 0.07) | 240 | 0.10 (0.08 to 0.11) | 270 | 0.11 (0.10 to 0.12) | 0.79 (0.67 to 0.92) |
| 75+ | max | 230 | 0.12 (0.10 to 0.13) | 210 | 0.10 (0.09 to 0.12) | 260 | 0.12 (0.11 to 0.14) | 150 | 0.07 (0.06 to 0.08) | 200 | 0.09 (0.08 to 0.10) | 260 | 0.11 (0.10 to 0.12) | 0.96 (0.80 to 1.14) |
| | min | 160 | 0.08 (0.07 to 0.09) | 140 | 0.07 (0.06 to 0.08) | 120 | 0.06 (0.05 to 0.07) | 100 | 0.05 (0.04 to 0.06) | 130 | 0.06 (0.05 to 0.07) | 180 | 0.08 (0.07 to 0.09) | 0.97 (0.78 to 1.19) |

**Table S6.** The max and min count and rate per 1,000 patients with a phenoxymethylpenicillin prescription and a record of scarlet fever, sore throat/tonsillitis, or iGAS up to 14 days prior or 7 days after the prescribing event for each season (September through August), by region. Counts have been rounded to the nearest 10 and the rate computed with the rounded number.

| | gas_year | 2017/18 | | 2018/19 | | 2019/20 | | 2020/21 | | 2021/22 | | 2022/23 | | 2023 v 2018 |
|---|---|---|---|---|---|---|---|---|---|---|---|---|---|---|
| | | Count | Rate (95% CI) | Count | Rate (95% CI) | Count | Rate (95% CI) | Count | Rate (95% CI) | Count | Rate (95% CI) | Count | Rate (95% CI) | Rate Ratio (95% CI) |
| group | type | | | | | | | | | | | | | |
| East | max | 12,210 | 2.21 (2.17 to 2.25) | 8,960 | 1.60 (1.56 to 1.63) | 9,530 | 1.67 (1.64 to 1.71) | 3,680 | 0.63 (0.61 to 0.65) | 7,960 | 1.37 (1.34 to 1.40) | 17,650 | 3.02 (2.98 to 3.07) | 1.37 (1.34 to 1.40) |
| | min | 5,030 | 0.90 (0.88 to 0.93) | 5,050 | 0.90 (0.88 to 0.93) | 1,850 | 0.32 (0.31 to 0.34) | 1,820 | 0.32 (0.30 to 0.33) | 4,990 | 0.86 (0.83 to 0.88) | 5,750 | 0.99 (0.96 to 1.01) | 1.10 (1.06 to 1.14) |
| East Midlands | max | 8,840 | 2.12 (2.08 to 2.17) | 6,750 | 1.59 (1.56 to 1.63) | 7,380 | 1.72 (1.68 to 1.76) | 2,980 | 0.69 (0.67 to 0.72) | 6,120 | 1.39 (1.35 to 1.42) | 13,530 | 3.05 (3.00 to 3.10) | 1.44 (1.40 to 1.48) |
| | min | 3,720 | 0.89 (0.86 to 0.92) | 3,760 | 0.90 (0.87 to 0.92) | 1,490 | 0.35 (0.33 to 0.36) | 1,560 | 0.36 (0.34 to 0.38) | 4,120 | 0.95 (0.92 to 0.98) | 4,500 | 1.02 (0.99 to 1.05) | 1.15 (1.10 to 1.20) |
| London | max | 1,940 | 1.26 (1.20 to 1.31) | 1,650 | 1.04 (0.99 to 1.09) | 1,910 | 1.15 (1.10 to 1.20) | 750 | 0.43 (0.40 to 0.46) | 1,540 | 0.85 (0.81 to 0.90) | 2,830 | 1.55 (1.50 to 1.61) | 1.24 (1.17 to 1.31) |
| | min | 900 | 0.58 (0.54 to 0.61) | 1,000 | 0.61 (0.58 to 0.65) | 370 | 0.22 (0.20 to 0.24) | 400 | 0.24 (0.21 to 0.26) | 950 | 0.53 (0.50 to 0.56) | 1,180 | 0.65 (0.62 to 0.69) | 1.13 (1.04 to 1.24) |
| North East | max | 2,450 | 2.15 (2.07 to 2.24) | 1,870 | 1.63 (1.55 to 1.70) | 1,820 | 1.57 (1.49 to 1.64) | 790 | 0.68 (0.63 to 0.73) | 1,630 | 1.38 (1.32 to 1.45) | 3,160 | 2.66 (2.57 to 2.76) | 1.24 (1.17 to 1.31) |
| | min | 1,010 | 0.88 (0.83 to 0.94) | 940 | 0.81 (0.76 to 0.87) | 430 | 0.37 (0.34 to 0.41) | 440 | 0.38 (0.34 to 0.41) | 910 | 0.78 (0.73 to 0.83) | 1,050 | 0.89 (0.84 to 0.94) | 1.01 (0.92 to 1.10) |
| North West | max | 5,370 | 2.54 (2.47 to 2.61) | 3,790 | 1.77 (1.72 to 1.83) | 3,640 | 1.69 (1.64 to 1.75) | 1,780 | 0.82 (0.79 to 0.86) | 3,540 | 1.61 (1.56 to 1.67) | 7,740 | 3.51 (3.43 to 3.59) | 1.38 (1.33 to 1.43) |
| | min | 2,010 | 0.94 (0.90 to 0.99) | 2,040 | 0.95 (0.91 to 0.99) | 850 | 0.39 (0.37 to 0.42) | 930 | 0.43 (0.40 to 0.46) | 2,220 | 1.02 (0.97 to 1.06) | 2,550 | 1.16 (1.11 to 1.20) | 1.23 (1.16 to 1.30) |
| South East | max | 2,970 | 1.87 (1.81 to 1.94) | 2,000 | 1.25 (1.19 to 1.30) | 1,890 | 1.17 (1.11 to 1.22) | 850 | 0.52 (0.48 to 0.55) | 1,720 | 1.04 (0.99 to 1.09) | 3,770 | 2.28 (2.20 to 2.35) | 1.22 (1.16 to 1.27) |
| | min | 1,160 | 0.73 (0.69 to 0.77) | 1,130 | 0.71 (0.67 to 0.75) | 410 | 0.25 (0.23 to 0.28) | 410 | 0.25 (0.23 to 0.28) | 1,050 | 0.64 (0.60 to 0.68) | 1,260 | 0.76 (0.72 to 0.80) | 1.05 (0.97 to 1.14) |
| South West | max | 5,100 | 1.55 (1.51 to 1.59) | 3,470 | 1.03 (1.00 to 1.07) | 3,800 | 1.11 (1.08 to 1.15) | 1,500 | 0.43 (0.41 to 0.45) | 3,460 | 0.98 (0.95 to 1.02) | 7,480 | 2.12 (2.07 to 2.17) | 1.37 (1.32 to 1.42) |
| | min | 2,220 | 0.67 (0.64 to 0.70) | 1,900 | 0.57 (0.55 to 0.60) | 740 | 0.22 (0.20 to 0.23) | 830 | 0.24 (0.22 to 0.26) | 1,990 | 0.57 (0.55 to 0.60) | 2,550 | 0.72 (0.70 to 0.75) | 1.08 (1.02 to 1.15) |
| West Midlands | max | 2,160 | 2.16 (2.07 to 2.25) | 1,760 | 1.73 (1.65 to 1.81) | 1,740 | 1.70 (1.62 to 1.78) | 890 | 0.87 (0.81 to 0.93) | 1,500 | 1.46 (1.39 to 1.53) | 3,120 | 3.02 (2.92 to 3.13) | 1.40 (1.33 to 1.48) |
| | min | 900 | 0.89 (0.83 to 0.95) | 920 | 0.90 (0.84 to 0.96) | 420 | 0.41 (0.37 to 0.45) | 400 | 0.39 (0.35 to 0.43) | 920 | 0.90 (0.84 to 0.95) | 1,130 | 1.10 (1.03 to 1.16) | 1.23 (1.13 to 1.34) |
| Yorkshire and The Humber | max | 7,920 | 2.25 (2.20 to 2.30) | 5,940 | 1.67 (1.63 to 1.71) | 6,590 | 1.84 (1.79 to 1.88) | 2,750 | 0.76 (0.73 to 0.78) | 5,950 | 1.62 (1.58 to 1.66) | 11,820 | 3.20 (3.14 to 3.26) | 1.42 (1.38 to 1.46) |
| | min | 3,540 | 1.00 (0.97 to 1.03) | 3,560 | 1.00 (0.96 to 1.03) | 1,370 | 0.38 (0.36 to 0.40) | 1,350 | 0.37 (0.35 to 0.39) | 3,690 | 1.01 (0.98 to 1.04) | 4,240 | 1.15 (1.12 to 1.19) | 1.15 (1.10 to 1.20) |

**Table S7.** The percent of prescriptions that had a GAS indication up to 14 days prior or 7 days after the prescribing event and the percent of clinical events that had a prescription up to 7 days prior and 14 days after the clinical event at the end of the study period (March 2023). Those with Invasive Strep A (iGAS) may be more likely to be treated in secondary care.

| | | Percent (95% CI) |
|---|---|---|
| **Medication with clinical any** | Phenoxymethylpenicillin | 40.6 (40.3 to 40.9) |
| | Clarithromycin | 11.3 (11.0 to 11.5) |
| | Erythromycin | 10.0 (9.5 to 10.4) |
| | Amoxicillin | 3.2 (3.1 to 3.2) |
| | Co_Amoxiclav | 1.3 (1.2 to 1.4) |
| | Cefalexin | 0.9 (0.8 to 1.0) |
| | Azithromycin | 0.7 (0.6 to 0.8) |
| | Flucloxacillin | 0.3 (0.2 to 0.3) |
| **Clinical with medication any** | Scarlet_Fever | 89.6 (88.6 to 90.6) |
| | Sore_Throat_Tonsillitis | 74.7 (74.4 to 75.0) |
| | Invasive_Strep_A | 28.6 (18.0 to 39.2) |

**Table S8.** Count of items relating to Group A strep treatment dispensed under Serious Shortage Protocol according to Freedom of Information (FOI) requests 30959 (covering Oct 2019 to Oct 2022) and 01234 (covering Nov 2022 to Apr 2023). 4 cefaclor items have been excluded as cefaclor was not included in this analysis. Code can be found https://github.com/opensafely/strepa_scarlet/blob/main/analysis/ssp-analysis/SSP-antibiotics.ipynb

| Antibiotic | Items |
|---|---|
| Amoxicillin | 20,320 |
| Phenoxymethylpenicillin | 11,875 |
| Flucloxacillin | 1,553 |
| Clarithromycin | 747 |
| Cefalexin | 473 |
| Erythromycin | 288 |
| Co-amoxiclav | 190 |
| Azithromycin | 12 |
| Total | 35,458 |
